# Distinct Peri-pubertal Growth Phenotypes Among Vietnamese Urban Children: A Functional Clustering Analysis of 2018-2025 School Health Data

**DOI:** 10.64898/2026.09.05.26362315

**Authors:** Nhan T. Ho

## Abstract

**Background:** Child growth is usually summarized by an average trajectory, an approach that can hide clinically meaningful heterogeneity between children. This study aims to identify various growth phenotypes in Vietnamese children.

**Methods:** This study analyzed retrospective longitudinal anthropometric data from annual school health check of children in Hanoi, Hochiminh, and Haiphong, Vietnam from 2018 to 2025. Children with at least 4 visits spanning at least 3 years were included (8,612 boys and 6,107 girls). Height for age trajectory was summarized with functional principal component scores and combined with SITAR derived random effects for growth size, pubertal timing, and tempo, then clustered with sex specific Gaussian mixture models. We evaluated phenotype associations with prepubertal BMI category, cluster stability, and whether phenotype improved prediction of late adolescent height beyond baseline prepubertal variables.

**Results:** Ten growth phenotypes in boys and twelve in girls were identified. Phenotype membership was associated with prepubertal BMI category in both sexes. A tall, accelerated growth phenotype in boys (4.2% of boys) had the highest prepubertal obesity prevalence (53.5%) and strongest obesity association in external validation. The girls’ phenotype solution was more sensitive to visit based eligibility criteria than the boys’ solution. Adding phenotype to a baseline model improved prediction of late adolescent height in both sexes, more strongly in boys.

**Conclusions:** Combining functional and SITAR based clustering identified clinically distinguishable growth phenotypes in Vietnamese children linked to prepubertal BMI and late adolescent height. Sex specific validation is needed before these phenotypes inform clinical application.

**Key messages:**

- Combining functional and SITAR based clustering identified 10 and 12 distinct growth phenotypes in Vietnamese boys and girls, linked to prepubertal BMI and improved prediction of late adolescent height.
- A tall, accelerated growth phenotype in boys carried the highest obesity prevalence in the cohort.
- Girls’ phenotypes were less stable, so sex specific validation is needed before clinical application.

## INTRODUCTION

Longitudinal studies of child growth are conventionally summarized by a single average trajectory plus a measure of variation around it, an approach that works well when a cohort grows in one broadly similar way but conceals meaningful structure when it does not ^1^. Over the past decade, a family of methods built to recover that hidden structure, often called growth phenotyping, has grown quickly, spanning growth mixture models, latent class growth analysis, longitudinal latent class analysis, functional principal component analysis, and more recently shape based and Bayesian nonparametric clustering ^1–3^.

Applied to real cohorts, these methods have consistently found that growth is not one story. Functional principal component analysis in South Indian birth cohorts identified height and weight trajectory patterns and the household and maternal factors linked to poor growth ^4,5^. In a Brazilian birth cohort, the same approach separated infants into growth archetypes labeled generally large, catch up, stunting, and faltering, with the stunting and faltering patterns predicting lower long term cognitive scores ^6^. Latent class mixed models in a South African birth cohort recovered three to five classes per anthropometric measure, strongly tied to rapid weight gain, stunting, and overweight ^7^. A Bayesian nonparametric model of joint height and weight trajectories in a Singaporean cohort found four growth subgroups ^2^, and feeding pattern clusters have been shown to separate distinct growth trajectories among Asian infants ^8^.

A recurring and somewhat uncomfortable finding in this literature is that the choice of clustering method changes the answer. One comparison found only 58% agreement between latent class mixed models and time series clustering for the same BMI trajectories, and agreement among different time series configurations alone ranged from 31 to 93% ^9^. The same body of work found that trajectory level features such as slope, tempo, and velocity, rather than the raw curve itself, could predict growth pattern membership with 82 to 89% accuracy ^9,10^, suggesting that features capturing the pace and timing of growth carry much of the signal that matters. This is close to the logic behind the SITAR growth curve model, which resolves each child’s height trajectory into interpretable random effects for size, timing, and tempo, and was designed specifically to capture the pubertal growth spurt ^11^.

Evidence from Vietnam sits mostly outside this methodological tradition. The multi country Young Lives cohort, following children from age 1 to 15 in Ethiopia, India, Peru, and Vietnam, remains the main source of longitudinal growth data from the region. Vietnamese children in this cohort were taller at age one and grew faster than their Ethiopian and Indian counterparts ^12^, and early socioeconomic disadvantage left a height gap that persisted through adolescence ^13^. These analyses relied on nonlinear mixed effects and latent growth curve models rather than functional or shape based clustering. Southeast Asian representation in the functional clustering literature is limited to the Singaporean cohort above and one Asian infant feeding study, and neither used functional principal component analysis ^2,8^. More broadly, growth phenotyping to date has concentrated on infancy and the first three to five years of life, built around outcomes like stunting and catch up growth. The years spanning later childhood into adolescence, when pubertal timing and tempo become the main source of individual variation, have not been studied this way, even in cohorts that follow children well into their teens ^12^.

This study addresses that gap. Using a large, multi-site Vietnamese school health cohort followed from early childhood through late adolescence, this study combine functional principal component scores of each child’s height for age trajectory with SITAR derived random effects for growth size, pubertal timing, and tempo, fit separately for boys and girls, to identify data driven growth phenotypes spanning the pubertal transition. This study describes these phenotypes, test their association with prepubertal BMI category and other cohort characteristics, and evaluate whether phenotype membership improves prediction of late adolescent height and persistent overweight or obesity beyond information already available before puberty begins.

## METHODS

### Study Population and Design

This study performed retrospective analysis of a longitudinal annual school health check program for children attending a private school system across three cities in Vietnam, Hanoi, Ho Chi Minh City, and Haiphong, from 2018 to 2025. This study was approved by Vinmec Ethical Committee (approval number 0231/2024/CN/HDDD VMEC) with a waiver of individual informed consent as the study used de-identified routinely collected retrospective health data from school health examinations.

Height and weight were converted to age and sex specific z scores using the World Health Organization 2007 growth reference for school aged children and adolescents ^14^. Individual height curves had already been summarized in earlier work from this cohort using the SITAR mixed effects growth model ^11^, fit separately for boys, ages 6 to 18 years, and girls, ages 6 to 16 years. This paper reused those fitted models and their random effects rather than refitting them.

### Clustering Eligible Sample

A child was eligible for the clustering analysis if they had at least 4 visits spanning at least 3 years within their sex specific SITAR fitting window and had a corresponding SITAR random effect from the earlier model. Children who fell in the same age window but did not meet the visit count or age span requirement were kept separately for a baseline comparison of cohort characteristics between eligible and non eligible children.

### Functional Principal Component Analysis

Each eligible child’s height for age z score trajectory was summarized with functional principal component analysis, implemented with the fdapace package for R, a method built for the sparse and irregularly timed measurements typical of a school health visit schedule ^15^. This analysis was run separately for boys and girls, since growth curve shape differs by sex. The number of components kept was the smallest set explaining at least 90% of the variance in trajectory shape for that sex.

### Clustering Input and Model Based Clustering

Two representations of each child’s growth pattern were combined for clustering, the retained functional principal component scores from the height trajectory, and the child’s SITAR random effects for the size, timing, and intensity of the pubertal growth spurt. Both sets of variables were standardized within sex before combining, since pooling raw values would let sex differences alone drive the clustering rather than differences in growth pattern within each sex. Clustering used Gaussian finite mixture models fit separately by sex with the mclust package for R ^16^. The number of clusters and the covariance structure were chosen by the Bayesian information criterion ^17^, searched from 2 to 12 clusters, with the integrated completed likelihood computed in parallel as a secondary check ^18^. Mean silhouette width was calculated for the chosen solution as a further measure of cluster separation ^19^.

### Phenotype Naming, Stability, and Validation

Each resulting cluster was given a descriptive name based on its own mean profile of overall height for age level, pubertal timing, and pubertal intensity, rather than an arbitrary cluster number. Cluster stability was assessed with 200 bootstrap resamples of children, refitting the mixture model on each resample and comparing the resulting classification of the original sample with the original classification using the adjusted Rand index ^20^. External validation described how phenotype membership related to prepubertal BMI category, hospital site, and birth year, using multinomial logistic regression fit separately by sex ^21^, with birth year centered on its cohort mean so that the model intercept fell within the observed range of the data.

### Sensitivity Analyses and Outcome Linkage

A sensitivity analysis compared clustering solutions built at three visit count eligibility thresholds, 3, 4, and 5 visits, using functional principal component scores alone since SITAR random effects are not available for children with fewer than 4 visits. Late adolescent height for age z score, taken from each child’s last visit between 16 and 18.5 years, and persistent overweight or obesity status were compared across phenotypes with one way analysis of variance and Fisher’s exact test ^22^, respectively, the latter chosen over the chi square test because several phenotype groups were small enough to produce sparse cross tabulation cells. Whether phenotype membership improved prediction of late adolescent height for age z score beyond baseline prepubertal variables was tested with a likelihood ratio test comparing nested linear regression models, one with baseline variables alone and one adding phenotype. A parallel analysis assessed how well phenotype membership could be predicted from prepubertal information alone, using five fold cross validated multinomial logistic regression compared against a no information baseline. All analyses were conducted in R, version 4.5.1 ^23^, and a two sided p value below 0.05 was considered statistically significant throughout.

## RESULTS

### Cohort and Clustering-Eligible Sample

Of the children with at least one visit in the sex-specific SITAR age window, 8,612 boys and 6,107 girls met the clustering eligibility criterion of at least 4 visits spanning at least 3 years and had a corresponding SITAR random effect from the earlier fitted model. Children who did not meet this criterion had a slightly later mean birth year and a lower mean prepubertal BMIZ than eligible children in both sexes, and showed a different hospital mix, with a larger share from HHP (**Table 1**).

**Table 1.** Baseline characteristics of the clustering-eligible cohort compared with children who did not meet the eligibility criterion, by sex.

| Sex | Group | N | Mean birth year | Mean HAZ baseline | Mean BMIZ baseline | Obesity (%) | HHN (%) | HCP (%) | HHP (%) |
| --- | --- | --- | --- | --- | --- | --- | --- | --- | --- |
| Male | Not eligible | 435 | 2014.2 | 0.16 | 1.14 | 19.8 | 50.8 | 25.1 | 24.1 |
| Male | Clustering-eligible | 8612 | 2012.4 | 0.38 | 1.55 | 26.9 | 64.9 | 26.5 | 8.6 |
| Female | Not eligible | 337 | 2014.7 | -0.01 | 0.32 | 5.6 | 52.5 | 23.4 | 24.0 |
| Female | Clustering-eligible | 6107 | 2013.2 | 0.24 | 0.51 | 7.9 | 63.6 | 28.3 | 8.1 |

HAZ = height-for-age z-score (WHO 2007 reference). BMIZ = body mass index-for-age z-score (WHO 2007 reference), both taken at the earliest visit at or before the prepubertal cutoff (11 years in boys, 9 years in girls).
Clustering-eligible children had at least 4 visits spanning at least 3 years within the sex-specific SuperImposition by Translation And Rotation (SITAR) fitting window (6 to 18 years in boys, 6 to 16 years in girls) and a fitted SITAR random effect. Not-eligible children had visits within that window but did not meet the visit-count or age-span criterion.
HHN= Hanoi, HCP= Hochiminh city, and HHP= Haiphong are the three study sites.

### Cluster Number and Phenotype Characteristics

The Bayesian information criterion, searched from 2 to 12 candidate clusters, selected 10 phenotypes for boys and 12 for girls, with a variable volume, variable shape, variable orientation covariance structure favored at every solution examined (**Figure 1**). The boys’ selection curve leveled off by the upper part of the searched range, and the neighboring 9 and 11-cluster solutions gave BIC values close to the chosen 10-cluster solution, though still slightly favoring more clusters (**Supplementary Table S1**). The girls’ curve was still rising at 12 clusters, the edge of the searched range, so the girls’ cluster count should be read as provisional. Phenotype size varied widely within each sex, from 60 to 2,173 children in boys and from 83 to 1,511 in girls (**Table 2**). The single largest phenotype was a stable, close-to-average height-for-age trajectory in boys and a persistently tall trajectory in girls. Smaller phenotypes in both sexes captured more distinct patterns, including early-timing and late-timing variants of the persistently tall pattern, and a slow-tempo, late catch-up pattern that appeared only among the boys’ phenotypes (**Table 2**, **Figures 2**, **Figure 3**).

**Figure 1.**
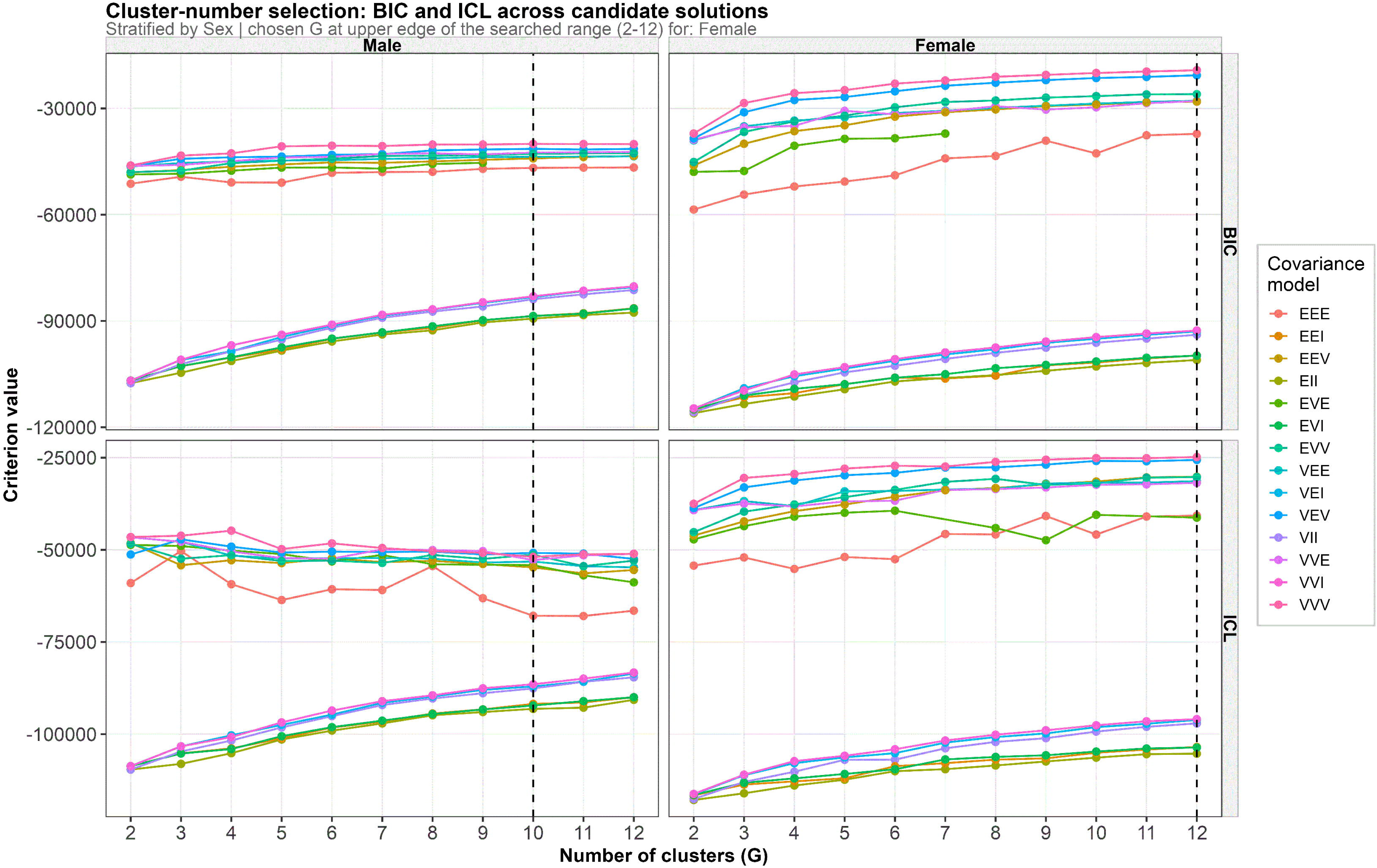
Cluster-number selection: BIC and ICL across candidate solutions, by sex. Bayesian information criterion (BIC, top row) and integrated completed likelihood (ICL, bottom row) for Gaussian mixture models fit to the combined functional-principal-component and SuperImposition by Translation And Rotation (SITAR) clustering input, across 2 to 12 candidate clusters (G) and thirteen candidate covariance structures (color), shown separately for boys (left column) and girls (right column). The dashed vertical line marks the solution chosen by BIC for each sex, G = 10 for boys and G = 12 for girls. The boys’ curves level off noticeably by the upper part of the searched range. The girls’ curves are still rising at G = 12, the upper edge of the search, so the girls’ cluster count should be treated as provisional pending a wider search in a future revision.

**Figure 2.**
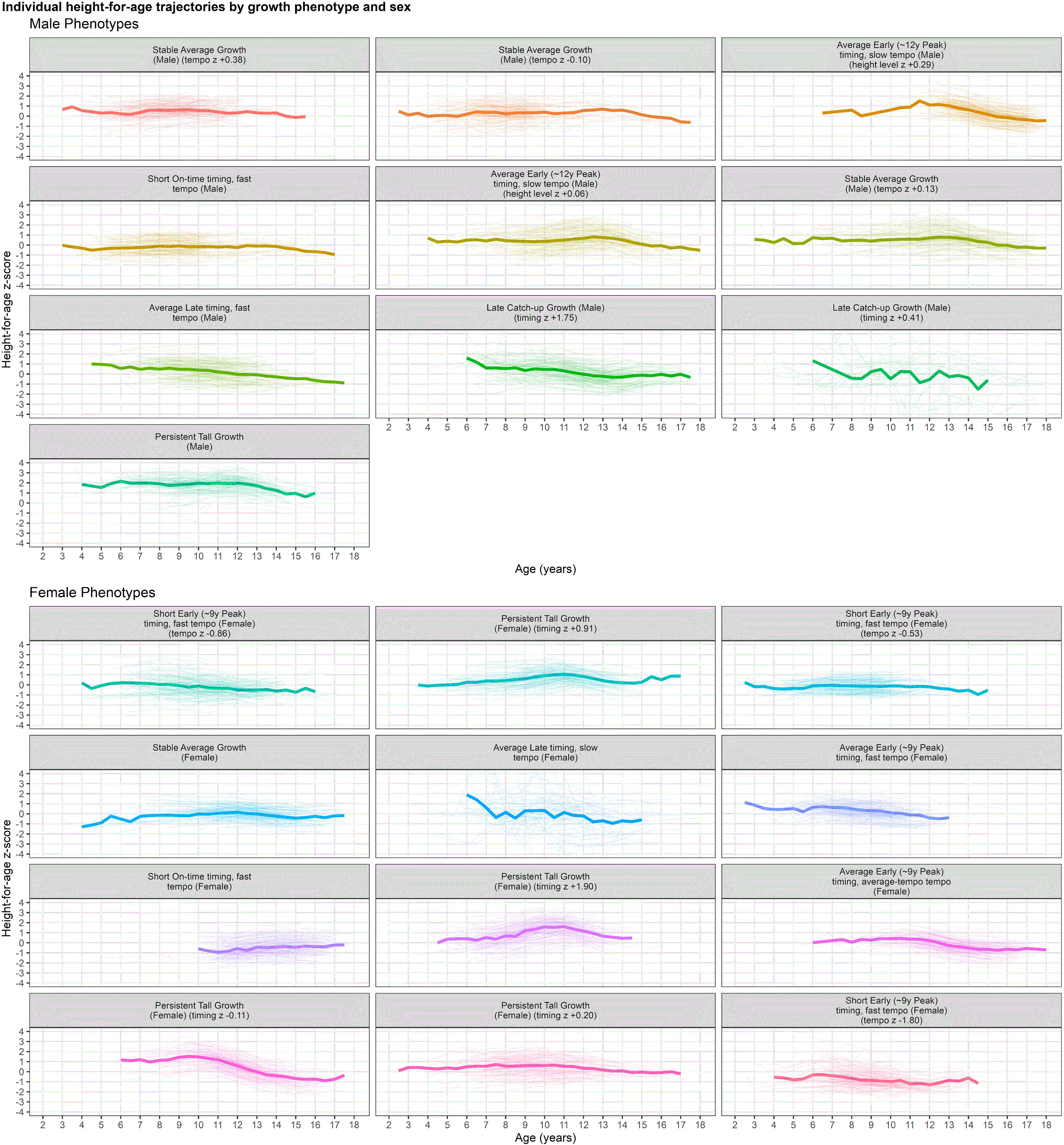
Individual height-for-age trajectories by growth phenotype and sex. Each panel shows a random subsample of up to 150 individual children’s height-for-age z-score trajectories (thin lines) for one growth phenotype, with the phenotype’s mean trajectory overlaid, a thick line computed from all children in that phenotype, not only the plotted subsample. Boys’ 10 phenotypes are shown in the top block, girls’ 12 phenotypes in the bottom block. Long phenotype names wrap onto multiple lines within each panel’s title rather than being cut off. Trajectories are height-for-age z-scores on the WHO 2007 growth reference. Phenotype names ending in a parenthetical qualifier (for example “(timing z +0.20)” or “(tempo z-0.53)”) denote two or more clusters that share the same qualitative description under the naming rule used here. The qualifier reports that cluster’s own value, to two decimals, on whichever of level, timing, or tempo separates the tied clusters the most, so the number is the cluster’s actual position on that axis rather than an arbitrary label.

**Figure 3.**
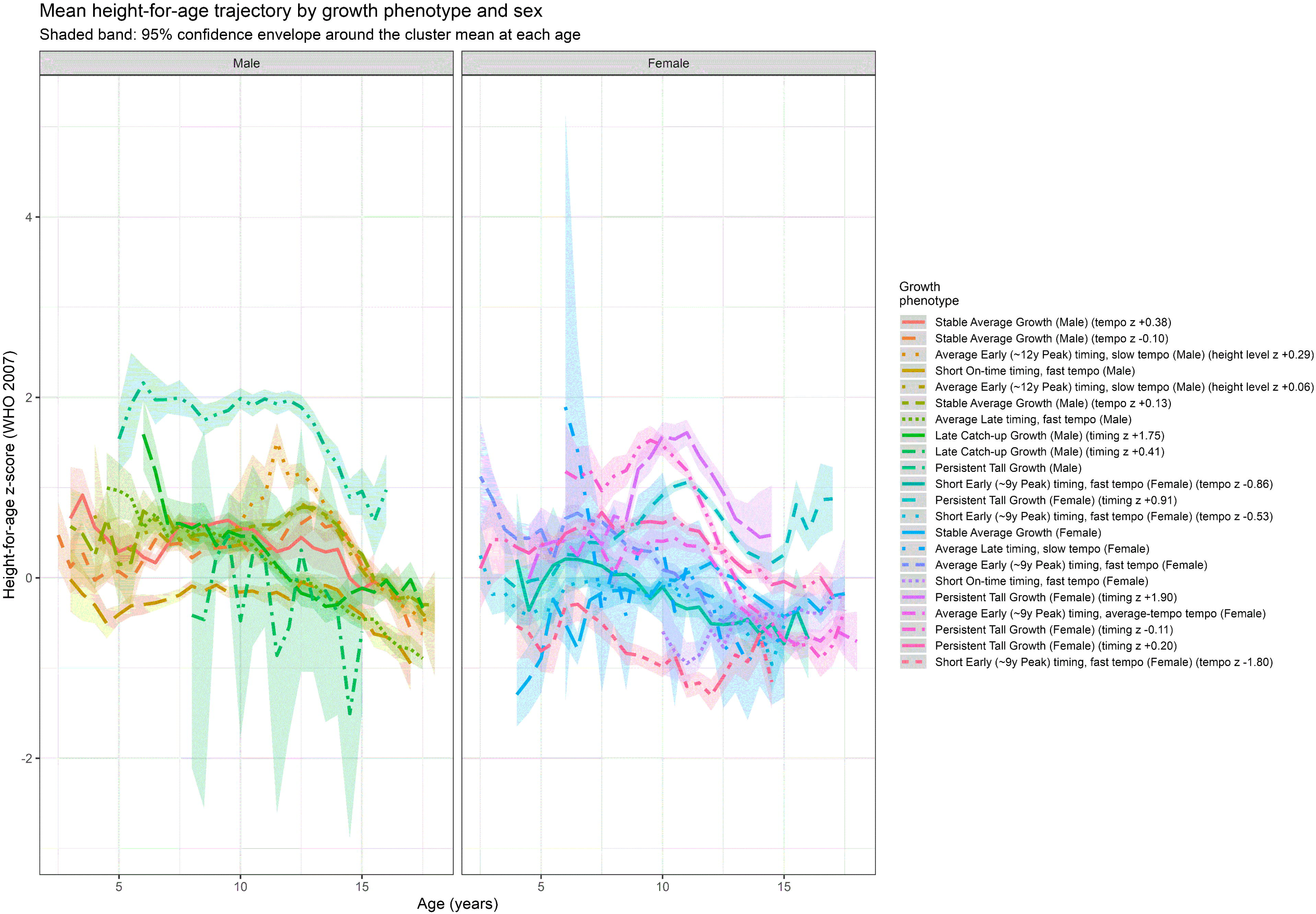
Mean height-for-age trajectory by growth phenotype and sex. Mean height-for-age z-score trajectory (line) with a 95 percent confidence envelope (shaded band) for each growth phenotype, plotted by age and faceted by sex (boys left, girls right). Values are computed within 6-month age bins that contain at least 10 children, and a phenotype is shown only if it has at least 6 such bins in an unbroken consecutive run, roughly 3 continuous years of coverage. Phenotypes that do not clear this bar are omitted from both the plot and the legend rather than left as an unexplained gap.

**Table 2.** Characteristics of the sex-specific growth phenotypes identified by combined functional and SITAR clustering.

| Sex | Growth phenotype | N | % of sex-specific cohort | Mean age at first visit (y) | Mean HAZ level (FPC1) | Mean SITAR timing (z) | Mean SITAR intensity (z) | Mean individual APHV (y) | Prepubertal obesity (%) |
| --- | --- | --- | --- | --- | --- | --- | --- | --- | --- |
| Male | Stable Average Growth (Male) (tempo z +0.13) | 2173 | 21.1 | 9.08 | 0.13 | -0.08 | 0.13 | 12.07 | 30.7 |
| Male | Average Early (~12y Peak) timing, slow tempo (Male) (height level z +0.06) | 1652 | 16.0 | 8.52 | 0.06 | -0.46 | 0.44 | 11.95 | 26.2 |
| Male | Short On-time timing, fast tempo (Male) | 1390 | 13.5 | 6.51 | -0.62 | 0.19 | -0.37 | 12.31 | 17.0 |
| Male | Average Late timing, fast tempo (Male) | 1342 | 13.0 | 8.07 | -0.25 | 0.77 | -0.7 | 12.6 | 32.4 |
| Male | Stable Average Growth (Male) (tempo z -0.10) | 1061 | 10.3 | 6.1 | -0.04 | 0.06 | -0.1 | 12.23 | 18.3 |
| Male | Stable Average Growth (Male) (tempo z +0.38) | 960 | 9.3 | 6.42 | 0.11 | -0.23 | 0.38 | 12.08 | 24.5 |
| Male | Average Early (~12y Peak) timing, slow tempo (Male) (height level z +0.29) | 886 | 8.6 | 9.01 | 0.29 | -0.48 | 0.35 | 11.83 | 31.7 |
| Male | Persistent Tall Growth (Male) | 438 | 4.2 | 8.19 | 1.49 | -0.99 | 1.05 | 11.68 | 53.5 |
| Male | Late Catch-up Growth (Male) (timing z +1.75) | 361 | 3.5 | 8.29 | -0.34 | 1.75 | -1.51 | 13.1 | 27.1 |
| Male | Late Catch-up Growth (Male) (timing z +0.41) | 60 | 0.6 | 7.29 | -0.56 | 0.41 | -0.44 | 12.4 | 42.6 |
| Female | Persistent Tall Growth (Female) (timing z +0.91) | 1511 | 17.2 | 7.37 | 0.54 | 0.91 | 0.91 | 10.11 | 6.9 |
| Female | Persistent Tall Growth (Female) (timing z +0.20) | 1487 | 17.0 | 6.73 | 0.32 | 0.2 | 0.2 | 9.4 | 8.9 |
| Female | Stable Average Growth (Female) | 1127 | 12.9 | 7.74 | -0.29 | -0.0 | -0.23 | 9.27 | 9.4 |
| Female | Short Early (~9y Peak) timing, fast tempo (Female) (tempo z -0.53) | 1037 | 11.8 | 6.05 | -0.44 | -0.41 | -0.53 | 8.83 | 4.6 |
| Female | Average Early (~9y Peak) timing, average-tempo tempo (Female) | 859 | 9.8 | 8.26 | -0.09 | -0.34 | -0.02 | 8.93 | 6.7 |
| Female | Average Early (~9y Peak) timing, fast tempo (Female) | 694 | 7.9 | 6.12 | -0.0 | -0.81 | -0.83 | 8.45 | 9.5 |
| Female | Persistent Tall Growth (Female) (timing z -0.11) | 604 | 6.9 | 8.0 | 0.58 | -0.11 | 0.69 | 9.3 | 18.8 |
| Female | Short Early (~9y Peak) timing, fast tempo (Female) (tempo z -0.86) | 471 | 5.4 | 7.38 | -0.58 | -0.81 | -0.86 | 8.44 | 7.6 |
| Female | Short Early (~9y Peak) timing, fast tempo (Female) (tempo z -1.80) | 422 | 4.8 | 6.37 | -1.24 | -1.41 | -1.8 | 7.89 | 2.2 |
| Female | Persistent Tall Growth (Female) (timing z +1.90) | 278 | 3.2 | 6.73 | 0.89 | 1.9 | 1.95 | 11.03 | 10.8 |
| Female | Short On-time timing, fast tempo (Female) | 197 | 2.2 | 7.27 | -1.02 | -0.19 | -0.92 | 8.47 | 0.0 |
| Female | Average Late timing, slow tempo (Female) | 83 | 0.9 | 6.71 | -0.26 | 1.95 | 1.36 | 11.7 | 13.8 |

Clustering combined standardized functional principal component (FPC) scores from each child's height-for-age z-score trajectory with standardized SuperImposition by Translation And Rotation (SITAR) random effects (size, timing, intensity), fit separately by sex. FPC1 ("HAZ level") summarizes each child's overall height-for-age position across the trajectory. SITAR timing is centered so that negative values indicate earlier pubertal timing than the sex-specific average, and SITAR intensity is centered so that negative values indicate a faster, more intense pubertal growth spurt.
APHV = age at peak height velocity, estimated individually from each child's SITAR random effects. Hospital-level percentages are reported instead in another table, because the join used to compute them for this table did not carry hospital code through and returned missing values for every phenotype.
The number of clusters was chosen separately by sex using BIC within a search range of G = 2 to 12. Boys chose G = 10, girls chose G = 12. The girls' solution sits at the upper edge of the searched range and should be treated as provisional.
HAZ = height-for-age z-score (WHO 2007 reference).

### Notable Phenotype Profiles by Sex

Several individual phenotypes stand out as clinically informative once their size, hospital and BMI associations, and downstream outcomes are considered together (**Table 2**, **Table 3**, **Table 4**, **Figure 3**, **Figure 4**). In boys, Persistent Tall Growth was one of the smaller phenotypes, accounting for only 4.2% of the male cohort, but it carried by far the highest prepubertal obesity rate of any boys’ phenotype, 53.5%, and showed the strongest external validation signal in the whole male solution, with about six times the odds of obesity and nearly three times the odds of overweight relative to the reference phenotype. Among the smaller number of boys with a recorded late-adolescent visit, this same phenotype also had the highest mean late-adolescent HAZ. Read together, this looks like an obesity-linked, accelerated growth pattern in boys that tracks into greater final height rather than a purely constitutional tall phenotype. By contrast, Short On-time timing, fast tempo, the third largest boys’ phenotype at 13.5% of the cohort, had the lowest prepubertal obesity rate among the larger phenotypes, 17.0%, lower odds of both overweight and obesity, and a lower mean late-adolescent HAZ, consistent with a more typical, non-obesity-driven growth pattern. A third comparison worth noting involves the two Late Catch-up Growth phenotypes, which share a qualitative label but diverge sharply. The larger of the two, 361 boys, showed no significant association with BMI category or birth year. The smaller one, only 60 boys, showed close to three times the odds of obesity, and among the very few boys in this group with a late-adolescent visit, the lowest mean HAZ and complete persistence of overweight or obesity, though this last estimate rests on only 13 children and should be read with caution.

**Figure 4.**
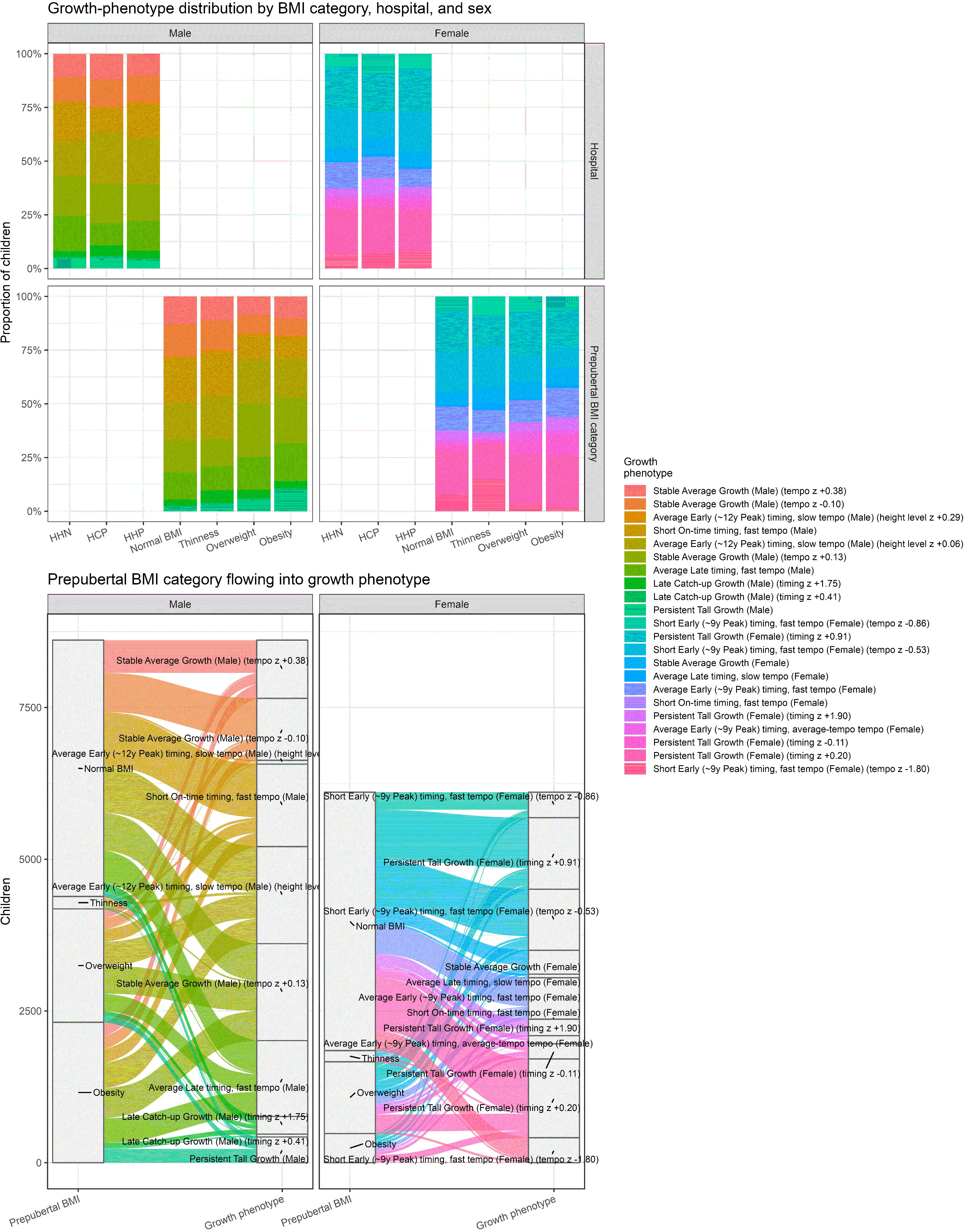
Growth-phenotype distribution by prepubertal BMI category, hospital, and sex, and the flow from BMI category into phenotype. Top: stacked bar charts showing the proportion of children in each growth phenotype (color), broken down by hospital (top row) and by prepubertal BMI category (bottom row), separately for boys (left) and girls (right). Bottom: alluvial diagram showing how children in each prepubertal BMI category, left-hand blocks, are distributed across growth phenotypes, right-hand blocks, separately for boys (left panel) and girls (right panel), with ribbon width proportional to the number of children following that path. Prepubertal BMI category is taken at the same baseline visit used throughout this study, 11 years or earlier in boys and 9 years or earlier in girls. With 22 phenotypes shown across both sexes, the alluvial diagram’s individual phenotype labels are dense and some overlap visually. More detailed source for exact phenotype-level numbers are presented in the tables.

**Table 3.**
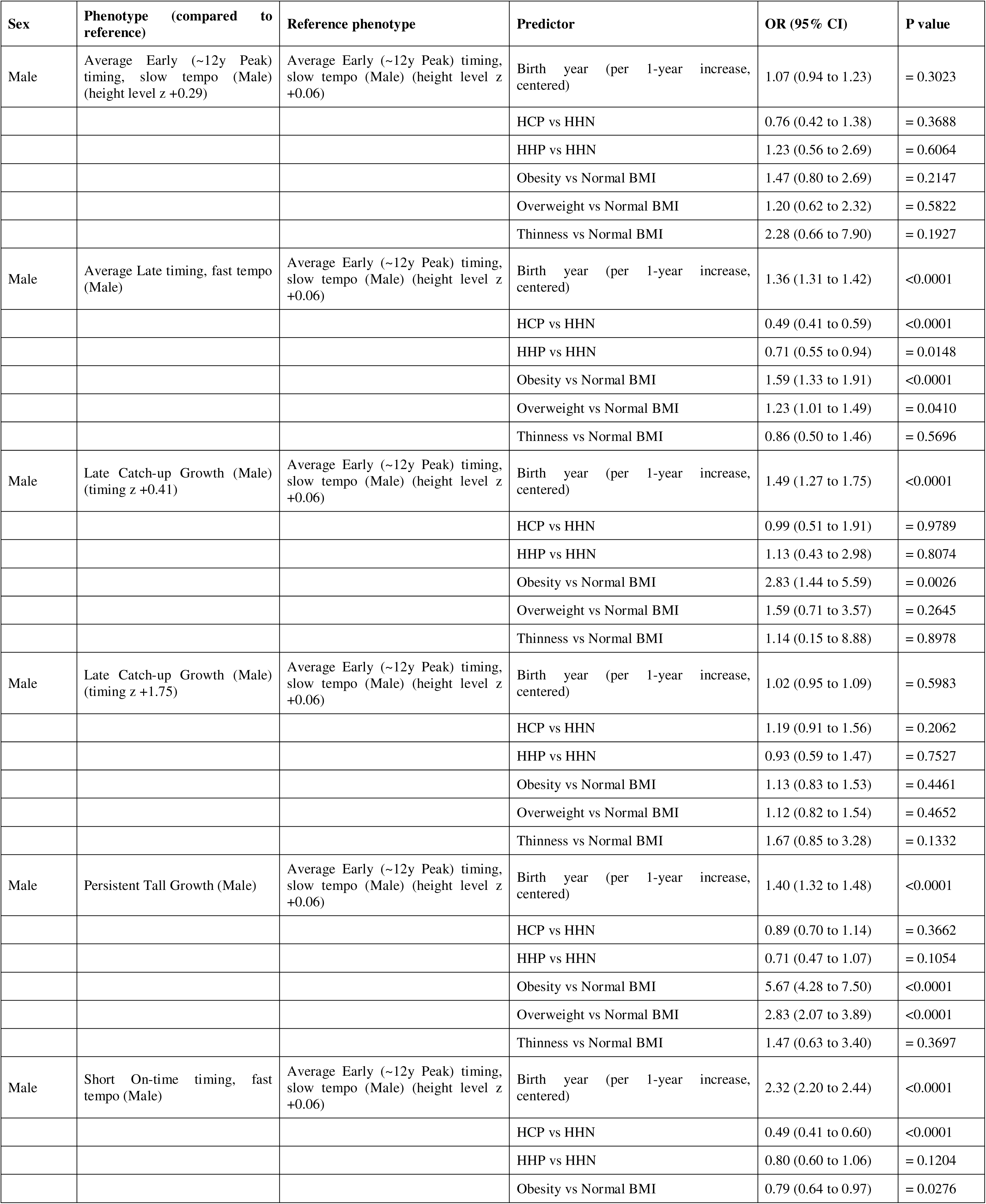
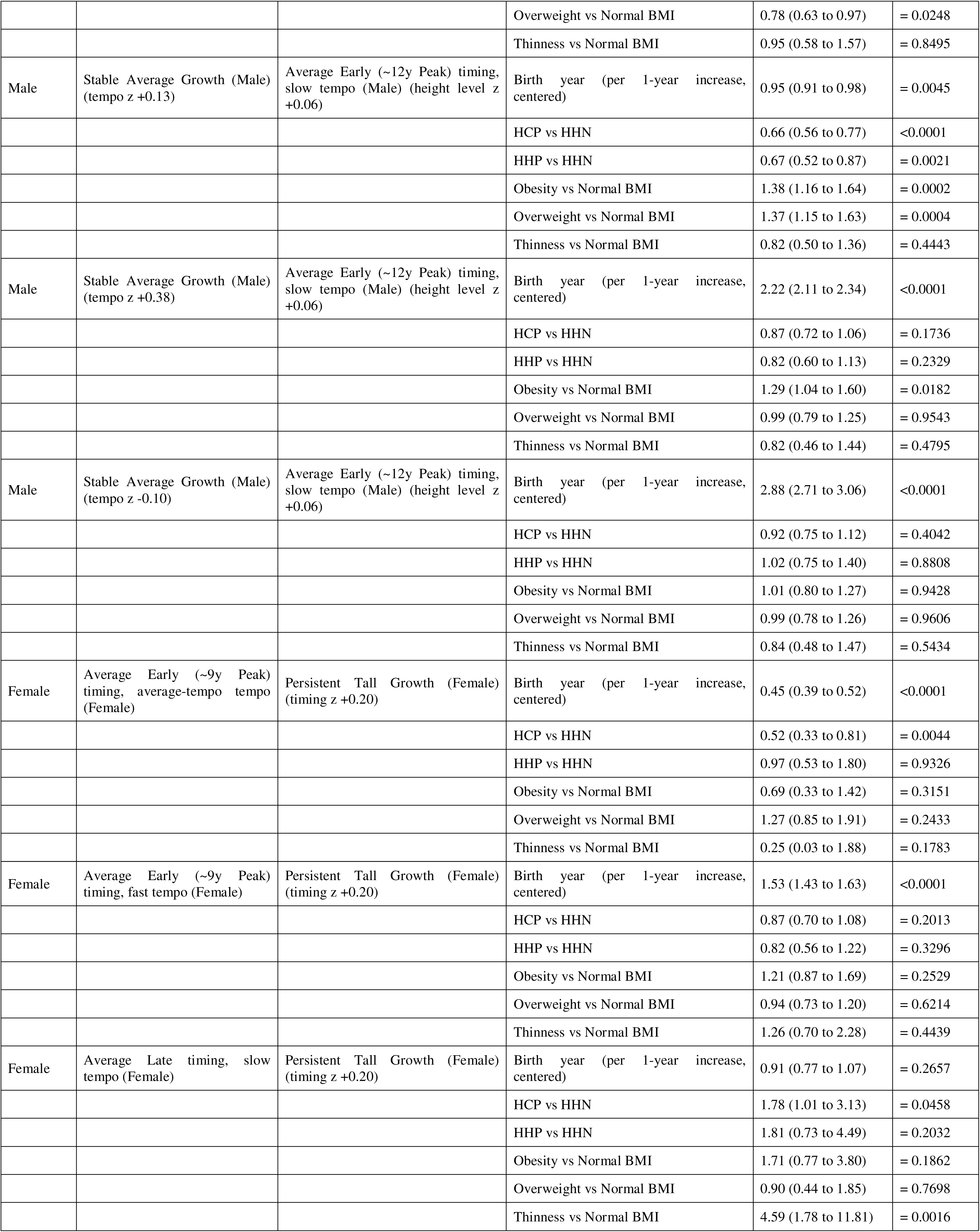
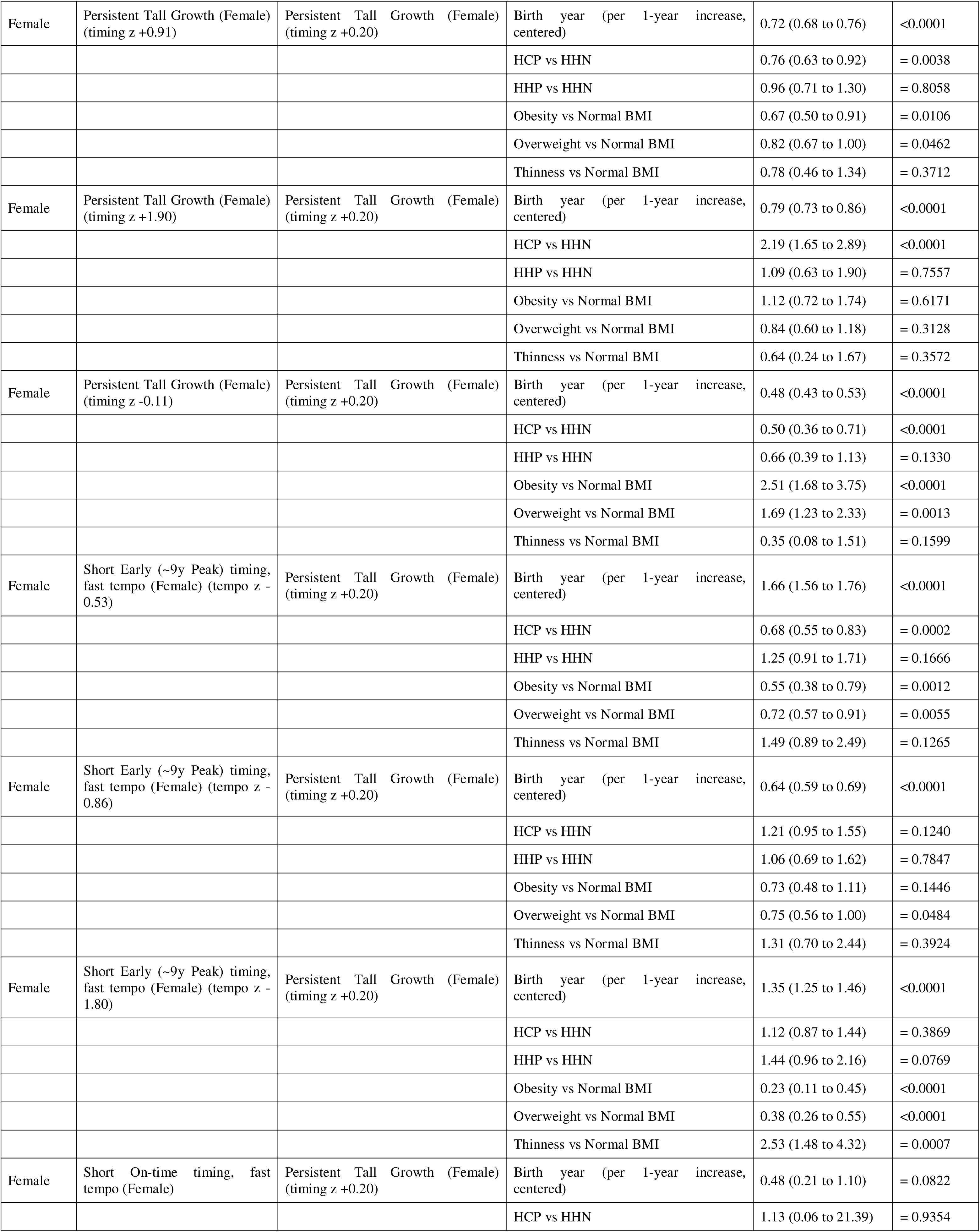
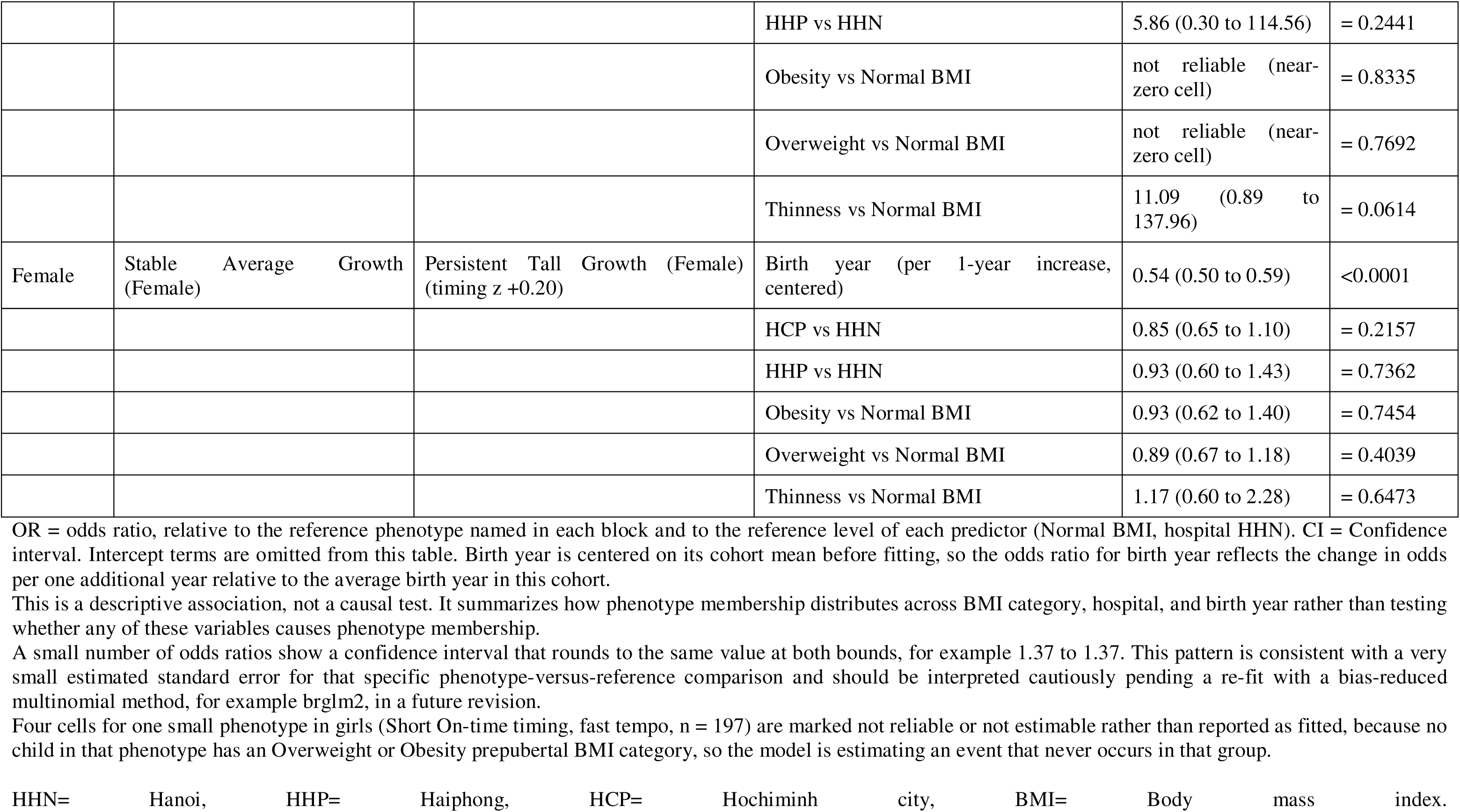
External validation: association of prepubertal BMI category, hospital, and birth year with growth-phenotype membership (multinomial logistic regression)

**Table 4.** Late-adolescent height-for-age z-score and persistent overweight/obesity status by growth phenotype.

| Sex | Growth phenotype | N with late-adolescent HAZ | Mean late-adolescent HAZ | SD | Mean individual APHV (y) | Persistent overweight/obesity (%) | ANOVA p (late-adolescent HAZ) | Fisher exact p (persistence) |
| --- | --- | --- | --- | --- | --- | --- | --- | --- |
| Male | Stable Average Growth (Male) (tempo z +0.13) | 744 | -0.23 | 0.72 | 12.03 | 37.7 | <0.0001 | not estimable |
| Male | Average Early (~12y Peak) timing, slow tempo (Male) (height level z +0.29) | 704 | -0.39 | 0.74 | 11.44 | 33.3 | <0.0001 | not estimable |
| Male | Average Early (~12y Peak) timing, slow tempo (Male) (height level z +0.06) | 210 | -0.24 | 0.82 | 12.29 | 30.3 | <0.0001 | not estimable |
| Male | Average Late timing, fast tempo (Male) | 102 | -0.79 | 0.41 | 12.73 | 19.4 | <0.0001 | not estimable |
| Male | Late Catch-up Growth (Male) (timing z +1.75) | 82 | -0.11 | 0.65 | 13.04 | 26.9 | <0.0001 | not estimable |
| Male | Stable Average Growth (Male) (tempo z -0.10) | 66 | -0.33 | 0.63 | 12.2 | 57.6 | <0.0001 | not estimable |
| Male | Short On-time timing, fast tempo (Male) | 60 | -0.8 | 0.55 | 12.36 | 22.9 | <0.0001 | not estimable |
| Male | Persistent Tall Growth (Male) | 18 | 0.63 | 0.73 | 11.7 | 40.0 | <0.0001 | not estimable |
| Male | Late Catch-up Growth (Male) (timing z +0.41) | 13 | -1.5 | 1.55 | 13.04 | 100.0 | <0.0001 | not estimable |
| Male | Stable Average Growth (Male) (tempo z +0.38) | 11 | -0.39 | 0.29 | 12.31 | 30.0 | <0.0001 | not estimable |
| Female | Average Early (~9y Peak) timing, average-tempo tempo (Female) | 321 | -0.66 | 0.7 | 9.01 | 25.0 | <0.0001 | = 0.8470 |
| Female | Stable Average Growth (Female) | 305 | -0.31 | 0.76 | 9.2 | 20.0 | <0.0001 | = 0.8470 |
| Female | Persistent Tall Growth (Female) (timing z -0.11) | 130 | -0.74 | 0.73 | 8.53 | 0.0 | <0.0001 | = 0.8470 |
| Female | Short On-time timing, fast tempo (Female) | 102 | -0.32 | 0.83 | NA | NA | <0.0001 | = 0.8470 |
| Female | Persistent Tall Growth (Female) (timing z +0.20) | 74 | -0.02 | 0.5 | 9.57 | 9.1 | <0.0001 | = 0.8470 |
| Female | Persistent Tall Growth (Female) (timing z +0.91) | 43 | 0.63 | 0.73 | 10.18 | 0.0 | <0.0001 | = 0.8470 |
| Female | Short Early (~9y Peak) timing, fast tempo (Female) (tempo z -0.86) | 17 | -0.51 | 0.94 | 7.81 | 0.0 | <0.0001 | = 0.8470 |
| Female | Average Late timing, slow tempo (Female) | 9 | -0.39 | 1.35 | NA | NA | <0.0001 | = 0.8470 |
| Female | Short Early (~9y Peak) timing, fast tempo (Female) (tempo z -0.53) | 7 | -0.63 | 0.46 | 9.12 | 0.0 | <0.0001 | = 0.8470 |
| Female | Average Early (~9y Peak) timing, fast tempo (Female) | 4 | 0.81 | 0.22 | 8.97 | 0.0 | <0.0001 | = 0.8470 |
| Female | Short Early (~9y Peak) timing, fast tempo (Female) (tempo z -1.80) | 3 | -0.6 | 0.35 | 8.48 | 0.0 | <0.0001 | = 0.8470 |
| Female | Persistent Tall Growth (Female) (timing z +1.90) | 1 | -0.38 | NA | 10.03 | 0.0 | <0.0001 | = 0.8470 |

Late-adolescent HAZ is each child's height-for-age z-score at their last recorded visit between 16 and 18.5 years of age. ANOVA p value tests whether mean late-adolescent HAZ differs across phenotypes within each sex.
Persistent overweight/obesity requires a child to have both a prepubertal BMI (body mass index) reading (Overweight or Obesity category) and a late-adolescent BMI reading (Overweight or Obesity category, age 16 to 18.5 years). The Fisher exact p value tests whether phenotype and persistent overweight/obesity status are associated within each sex, using one test per sex whose result is repeated on every row for that sex. This returns a real result for girls (p = 0.847, not significant). It still fails for boys and returns not estimable, most likely because the phenotype-by-persistence cross-tabulation is too sparse for the exact algorithm to complete even for boys' larger sample. Boys' percentages are still reported as descriptive information.
Sample sizes for this table are smaller because a late-adolescent visit, 16 to 18.5 years, was not available for every clustering-eligible child, and some of the smallest phenotype groups accordingly have very few children, as low as n = 1, so their percentages and mean APHV (where shown as NA for too few children) should be read as unstable.

In girls, Persistent Tall Growth split into several phenotypes that differed meaningfully from one another despite the shared label. The single largest phenotype in the entire female solution, accounting for 17.2% of girls, combined a relatively low prepubertal obesity rate of 6.9% with lower odds of obesity than the similarly tall reference phenotype, and the highest mean late-adolescent HAZ among girls with follow-up data. A smaller, more extreme variant of the same pattern, marked by earlier timing and higher intensity and comprising 3.2% of girls, carried a higher prepubertal obesity rate of 10.8% and was more strongly tied to hospital site, though its association with BMI category did not reach significance. At the other end, the most extreme Short Early, fast-tempo phenotype, 4.8% of girls, had the lowest prepubertal obesity rate of any female phenotype, 2.2%, roughly a quarter the odds of obesity and around a third the odds of overweight relative to the reference phenotype, alongside more than double the odds of thinness. One further phenotype deserves mention on clinical grounds rather than statistical ones. A small, on-time, fast-tempo group of 197 girls had no prepubertal overweight or obesity at all, so its association with BMI category could not be estimated. Whatever drives membership in this phenotype, it is not excess prepubertal weight, and it stands apart from every other girls’ phenotype in that respect.

### External Validation

Beyond the specific phenotypes described above, growth phenotype membership was associated with prepubertal BMI category, hospital site, and birth year across the broader set of phenotypes in both sexes (**Table 3**). Hospital site and birth year were associated with phenotype membership in most comparisons, consistent with the cohort’s multi-site, multi-year recruitment (**Table 3**, **Figure 4**). One small phenotype in girls, the on-time, fast-tempo group of 197 children noted above, could not be evaluated for its association with BMI category, since the corresponding coefficients had no informative variation to estimate from.

### Cluster Stability and Sensitivity Analyses

Across bootstrap resamples, the median adjusted Rand index comparing the original classification with each resample’s classification was 0.46 in boys and 0.50 in girls, indicating moderate rather than high stability in both sexes (**Supplementary Table S2**). Clustering built on functional principal component scores alone was essentially unchanged across the 3, 4, and 5-visit eligibility thresholds in boys, with an adjusted Rand index above 0.95 at both comparisons. In girls, the same comparison showed much lower agreement, an adjusted Rand index of 0.65 between the 3 and 4-visit thresholds and 0.16 between the 4 and 5-visit thresholds, indicating that the girls’ phenotype solution is considerably more sensitive to this choice than the boys’ solution (**Supplementary Table S3**).

### Late-Adolescent Outcomes and Predictive Value

Late-adolescent height-for-age z score differed significantly across phenotypes in both sexes (**Table 4**), with the highest values generally seen in phenotypes already identified as persistently tall and the lowest values concentrated among the smaller, less common phenotypes. Persistent overweight or obesity, present at both a prepubertal and a late-adolescent visit, also varied across phenotypes descriptively, though the Fisher exact test for this association returned a usable result only in girls, where it was not statistically significant, and could not be completed in boys.

Adding growth phenotype to a baseline model of prepubertal BMIZ, baseline height-for-age z score, sex, and hospital improved prediction of late-adolescent height-for-age z score in both sexes. In boys, the adjusted R-squared rose from 0.580 to 0.607, a statistically significant improvement by likelihood ratio test. In girls, the adjusted R-squared rose from 0.262 to 0.431, also statistically significant, though based on only 52 girls with complete baseline and late-adolescent data (**Table 5**).

**Table 5.** Does growth phenotype improve prediction of late-adolescent HAZ beyond baseline variables.

| Sex | Model | Adjusted R-squared | AIC | N | LRT p vs Model A |
| --- | --- | --- | --- | --- | --- |
| Male | A: baseline only | 0.58 | 820.1 | 599 | - |
| Male | B: baseline + phenotype | 0.607 | 789.1 | 599 | <0.0001 |
| Female | A: baseline only | 0.262 | 100.6 | 52 | - |
| Female | B: baseline + phenotype | 0.431 | 94.3 | 52 | = 0.0193 |

Model A predicts late-adolescent HAZ from prepubertal BMIZ, baseline HAZ, sex, and hospital. Model B adds growth phenotype to Model A. LRT p value is from a likelihood ratio test of Model B against Model A within each sex. HAZ = height-for-age z-score (WHO 2007 reference). BMIZ = body mass index-for-age z-score (WHO 2007 reference).
In boys, adding phenotype improves the adjusted R-squared from 0.580 to 0.607 and lowers the Akaike Information Criterion (AIC) by about 31 points, a statistically significant improvement, LRT p less than 0.0001. In girls, the same comparison also reaches significance, adjusted R-squared rising from 0.262 to 0.431, LRT p equal to 0.019, though the sample size for this analysis is still small, n equal to 52, and this should be read as a real but fragile signal pending replication in a larger late-adolescent follow-up sample.
The small girls' sample size for this analysis, n equal to 52, reflects how few girls have both complete baseline data and a recorded late-adolescent visit, and limits how much weight the girls' result in this table can carry.

Five-fold cross-validated prediction of phenotype membership from prepubertal BMIZ, baseline height-for-age z score, and hospital alone exceeded a no-information baseline by 7.0 percentage points in boys and 3.3 percentage points in girls, indicating that growth phenotype is only partly determined by information available before puberty begins (**Table 6**).

**Table 6.** Five-fold cross-validated accuracy of predicting growth phenotype from prepubertal information alone.

| Sex | CV folds | Mean accuracy (%) | No-information accuracy (%) | Improvement (percentage points) |
| --- | --- | --- | --- | --- |
| Male | 5 | 26.7 | 19.7 | 7.0 |
| Female | 5 | 24.2 | 20.9 | 3.3 |

Predictors are prepubertal BMIZ, baseline HAZ, and hospital, fit separately within each sex. No-information accuracy is the accuracy of always predicting the most common phenotype in the training fold, the baseline a useful model must beat. HAZ = height-for-age z-score (WHO 2007 reference). BMIZ = body mass index-for-age z-score (WHO 2007 reference).
Prepubertal information alone gives a modest improvement over the no-information baseline in both sexes, about 3.3 percentage points in girls and about 7.0 percentage points in boys, consistent with growth phenotype being only partly determined by information available before puberty begins.

## DISCUSSION

This study applied functional principal component analysis combined with SITAR derived pubertal random effects to a large Vietnamese school health cohort and identified 10 growth phenotypes in boys and 12 in girls, spanning childhood into late adolescence. Phenotype membership was associated with prepubertal BMI category in both sexes, and adding phenotype to a baseline prepubertal model improved prediction of late adolescent height, most clearly in boys. To our knowledge, this combination of a whole trajectory functional descriptor with SITAR’s pubertal size, timing, and tempo random effects has not been applied together before, and rarely at all in a Southeast Asian population.

The moderate cluster stability observed here, a median adjusted Rand index of 0.46 in boys and 0.50 in girls, fits a broader pattern already reported in the growth phenotyping literature, where different clustering methods and even different resamples of the same data can meaningfully change which groups emerge ^9^. Most published functional principal component work on child growth has concentrated on infancy and the first several years of life, often around a single anthropometric measure and outcomes like stunting or catch up growth ^4,6^. Extending this approach into the pubertal years, where timing and tempo rather than absolute size become the dominant source of individual variation, appears to be new ground rather than a straightforward replication of existing methods.

The clearest clinical signal was a boys’ phenotype defined by tall stature and accelerated growth that carried both the highest prepubertal obesity prevalence of any boys’ phenotype and the strongest obesity association in external validation, echoing a South African birth cohort where latent growth classes were similarly tied to rapid weight gain and overweight ^7^. A more cautionary finding involved the two Late Catch-up Growth phenotypes in boys, which shared a qualitative label but differed sharply in their BMI association and later outcomes. Growth patterns that look similar enough to earn the same descriptive name can still carry very different clinical meaning, consistent with earlier work showing that different clustering choices applied to the same trajectories can disagree substantially on group membership ^9,10^.

A finding with no direct precedent in the reviewed literature was the marked sex asymmetry in phenotype robustness. The girls’ solution was far more sensitive to the visit count eligibility threshold than the boys’, and the girls’ cluster count sat at the edge of the searched range rather than settling on a clear interior optimum. Girls in this cohort enter and complete the pubertal growth spurt earlier and over a narrower age window than boys, which may leave fewer usable visits per child within a fixed eligibility rule and make the phenotype solution more sensitive to exactly which children and visits are retained. This is a plausible explanation rather than a tested one, and it points to a methodological question future work in this area should address directly.

Several limitations temper these findings. The girls’ cluster count was not fully resolved within the searched range and should be treated as provisional. Stability was moderate rather than high in both sexes, so the phenotypes described here are best read as a reasonable data driven summary rather than a fixed taxonomy. The exact test linking phenotype to persistent overweight or obesity could not be completed for boys, and the predictive value analysis in girls rested on only 52 children with complete late adolescent follow up. This is a single country, three city school cohort, and whether these phenotypes and their BMI associations hold in other Vietnamese or Southeast Asian populations remains untested.

For clinical practice, the tall, fast growing, high obesity phenotype identified in boys suggests that rapid or early pubertal progression paired with excess prepubertal weight is worth treating as a signal for closer metabolic follow up, rather than assuming tall stature in childhood is reassuring on its own. For public health, the consistent link between phenotype and BMI category in both sexes, together with the modest ability of prepubertal information alone to predict later phenotype, supports continued growth and weight surveillance through the pubertal transition rather than concentrating monitoring effort on the prepubertal years alone. More broadly, this study extends growth phenotyping into an age range and population where it has rarely been used, and the sex difference in phenotype stability found here suggests that future studies should test and report robustness separately for boys and girls rather than assuming a method that performs well in one sex will perform equally well in the other.

In conclusion, combining functional and SITAR based clustering identified biologically interpretable growth phenotypes in this Vietnamese cohort that were tied to prepubertal BMI, captured sex specific patterns of pubertal timing and tempo, and improved prediction of late adolescent height beyond routinely collected prepubertal variables.

## Data Availability

R codes are available from the corresponding author upon reasonable request. Individual patient-level data cannot be shared due to applicable privacy regulations and the terms of the institutional ethics approval.

## DECLARATION

### Contributor’s statement

Nhan Thi Ho did conceptualization, data curation, formal analysis, investigation, methodology, project administration, resources, software, supervision, validation, visualization, writing original draft, and writing review & editing.

### Conflict of intertest

The author states that there is no conflict of interest.

### Use of Artificial Intelligence

The author performed all original research work regarding scientific content, analyses, interpretations and manuscript writing. The author used AI-assisted tools for language editing and grammar checking during manuscript preparation.

## REFERENCES

1. Herle M, Micali N, Abdulkadir M, et al. Identifying typical trajectories in longitudinal data: modelling strategies and interpretations. Eur J Epidemiol. 2020;35:205–222. doi:10.1007/s10654-020-00615-6

2. Beraha M, Guglielmi A, Quintana F, De Iorio M, Eriksson J, Yap F. Childhood obesity in Singapore: A Bayesian nonparametric approach. Stat Modelling. 2023;24:541–560. doi:10.1177/1471082x231185892

3. López-Domínguez L, Bassani D, Bourdon C, et al. A novel shape-based approach to identify gestational age-adjusted growth patterns from birth to 11 years of age. Sci Rep. 2023;13. doi:10.1038/s41598-023-28485-4

4. Karuppusami R, Antonisamy B, Premkumar P. Functional principal component analysis for identifying the child growth pattern using longitudinal birth cohort data. BMC Med Res Methodol. 2022;22. doi:10.1186/s12874-022-01566-0

5. Karuppusami R, Belavendra A, Premkumar P. Identifying longitudinal child growth curve pattern using Functional Principal Component Analysis from birth cohort study in South India. Published online 2021. doi:10.21203/rs.3.rs-225326/v1

6. Han K, Hadjipantelis P, Wang JL, et al. Functional principal component analysis for identifying multivariate patterns and archetypes of growth, and their association with long-term cognitive development. PLoS One. 2018;13. doi:10.1371/journal.pone.0207073

7. Van Biljon N, Lake M, Goddard L, Botha M, Zar H, Little F. Latent classes of anthropometric growth in early childhood using uni- and multivariate approaches in a South African birth cohort. PLoS One. 2025;20. doi:10.1371/journal.pone.0319237

8. Van Der Merwe L, Mulder K, Van Oudenhoven F, Shek L, Teoh O, Pang WW. Mixed Milk Feeding Patterns and Growth Outcomes During the First Year of Life in Asian Infants: Application of Predefined Feeding Clusters to Test Associations. Curr Dev Nutr. 2025;9. doi:10.1016/j.cdnut.2025.107565

9. Massara P, Keown-Stoneman C, Erdman L, et al. Identifying longitudinal-growth patterns from infancy to childhood: a study comparing multiple clustering techniques. Int J Epidemiol. Published online 2021. doi:10.1093/ije/dyab021

10. Massara P, López-Domínguez L, Bourdon C, et al. A novel systematic pipeline for increased predictability and explainability of growth patterns in children using trajectory features. Int J Med Inform. 2023;177:105143. doi:10.2139/ssrn.4359177

11. Cole TJ, Donaldson MDC, Ben-shlomo Y. SITAR-a useful instrument for growth curve analysis. Int J Epidemiol. 2010;39(6). doi:10.1093/ije/dyq115

12. Wake SK, Zewotir T, Muluneh EK. Studying latent change process in height growth of children in Ethiopia, India, Peru and Vietnam. BMC Pediatr. 2022;22(1). doi:10.1186/s12887-022-03269-3

13. Aizawa T. Trajectory of inequality of opportunity in child height growth: Evidence from the Young Lives study. Demogr Res. Published online 2020. doi:10.4054/demres.2020.42.7

14. De Onis M, Onyango AW, Borghi E, Siyam A, Nishida C, Siekmann J. Development of a WHO growth reference for school-aged children and adolescents. Bull World Health Organ. 2007;85(9). doi:10.2471/BLT.07.043497

15. Yao F, Müller HG, Wang JL. Functional data analysis for sparse longitudinal data. J Am Stat Assoc. 2005;100(470). doi:10.1198/016214504000001745

16. Scrucca L, Fop M, Murphy TB, Raftery AE. Mclust 5: Clustering, classification and density estimation using Gaussian finite mixture models. R Journal. 2016;8(1). doi:10.32614/rj-2016-021

17. Schwarz G. Estimating the Dimension of a Model. The Annals of Statistics. 2007;6(2). doi:10.1214/aos/1176344136

18. Biernacki C, Celeux G, Govaert G. Assessing a mixture model for clustering with the integrated completed likelihood. IEEE Trans Pattern Anal Mach Intell. 2000;22(7). doi:10.1109/34.865189

19. Rousseeuw PJ. Silhouettes: A graphical aid to the interpretation and validation of cluster analysis. J Comput Appl Math. 1987;20(C). doi:10.1016/0377-0427(87)90125-7

20. Hubert L, Arabie P. Comparing partitions. J Classif. 1985;2(1). doi:10.1007/BF01908075

21. Venables WN, Ripley BD. Modern Applied Statistics with S. Fourth. Springer; 2002. https://www.stats.ox.ac.uk/pub/MASS4/

22. Fisher RA. On the Interpretation of χ 2 from Contingency Tables, and the Calculation of P. Journal of the Royal Statistical Society. 1922;85(1). doi:10.2307/2340521

23. R Core Team. R: A Language and Environment for Statistical Computing. R Foundation for Statistical Computing, Vienna, Austria. Preprint posted online 2016. doi:10.1017/CBO9781107415324.004

