## Supplementary Tables for "Distinct Peri-pubertal Growth Phenotypes Among Vietnamese Urban Children: A Functional Clustering Analysis of 2018-2025 School Health Data"

**Supplementary Table S1. Neighboring cluster-number solutions (G-1, the chosen G, and G+1 where fit) compared by BIC and resulting cluster sizes.**

| **Sex** | **Solution** | **G** | **Covariance model** | **BIC** | **Cluster sizes (n)** |
| --- | --- | --- | --- | --- | --- |
| Male | G-1 | 9 | VVV | -40069.4 | 1407, 1325, 794, 2409, 992, 2151, 305, 59, 881 |
| Male | G (chosen) | 10 | VVV | -40019.5 | 960, 1061, 886, 1390, 1652, 2173, 1342, 361, 60, 438 |
| Male | G+1 | 11 | VVV | -39993.4 | 1588, 127, 856, 923, 929, 361, 2200, 875, 58, 1040, 1366 |
| Female | G-1 | 11 | VVV | -19570.4 | 961, 2055, 289, 539, 100, 1463, 605, 992, 1199, 353, 214 |
| Female | G (chosen) | 12 | VVV | -19219.0 | 471, 1511, 1037, 1127, 83, 694, 197, 278, 859, 604, 1487, 422 |

Boys' chosen solution, G = 10, sits inside the searched range (2 to 12), and both neighboring solutions were fit for comparison. BIC continues to improve slightly from G = 9 through G = 11, so the boys' solution, while much better resolved than in earlier, narrower searches, is still not a sharply peaked optimum.

Girls' G+1 solution is not shown because the chosen solution, G = 12, was already at the upper edge of the searched range, so a G = 13 solution was not fit. This is the boundary issue described on the cover page and it remains specific to girls in this run.

VVV indicates a Gaussian mixture model with variable volume, variable shape, and variable orientation across clusters, the covariance model selected by BIC for every solution shown.

**Supplementary Table S2. Cluster stability: adjusted Rand index between the original clustering and 200 bootstrap-resampled refits, by sex**

| **Sex** | **Bootstrap reps** | **Valid reps** | **Median ARI** | **25th pct ARI** | **75th pct ARI** | **Minimum ARI** |
| --- | --- | --- | --- | --- | --- | --- |
| Male | 200 | 200 | 0.455 | 0.393 | 0.517 | 0.279 |
| Female | 200 | 200 | 0.496 | 0.415 | 0.572 | 0.292 |

Each bootstrap replicate resamples children with replacement, refits the same mixture model on the resampled data, classifies the original sample under that refit, and compares the result with the original classification using the adjusted Rand index, where 1.0 means identical and 0 means no better than chance.

Median adjusted Rand index is 0.46 in boys and 0.50 in girls. This indicates moderate, not high, stability and should be reported as a limitation of the clustering solution in both sexes, not as strong evidence of sharply distinct, well-separated phenotypes.

**Supplementary Table S3. Sensitivity of the functional-principal-component-only clustering solution to the visit-count eligibility threshold, by sex**

| **Sex** | **Visit-count comparison** | **ARI** | **Children in both solutions** |
| --- | --- | --- | --- |
| Male | 3 vs 4 visits | 0.956 | 10323 |
| Male | 4 vs 5 visits | 0.955 | 5431 |
| Female | 3 vs 4 visits | 0.654 | 8770 |
| Female | 4 vs 5 visits | 0.163 | 4411 |

This sensitivity analysis uses functional principal component scores alone, without SuperImposition by Translation And Rotation (SITAR) random effects, because SITAR itself requires at least 4 visits and so cannot be refit at the 3-visit threshold without a separate SITAR model.

Boys' clustering is essentially unchanged across the 3, 4, and 5-visit thresholds, adjusted Rand index above 0.95 at both comparisons. Girls' clustering changes substantially, adjusted Rand index of 0.65 comparing the 3 and 4-visit thresholds and 0.16 comparing the 4 and 5-visit thresholds. The girls' phenotype solution remains considerably more dependent on which eligibility threshold is chosen than the boys' solution, and this should be discussed as a limitation specific to the girls' phenotypes.
